# Does combining the STarT Back Tool with a polygenic risk score for chronic low back pain improve prediction of work disability over two years?

**DOI:** 10.1101/2025.09.09.25335394

**Authors:** Roger Compte, Maryam Kazemi Naeini, Eveliina Heikkala, Terence McSweeney, Jaro Karppinen, Frances MK Williams

**Affiliations:** Department of Twin Research and Genetic Epidemiology, School of Life Course and Population Sciences, King’s College London, London, UK; Research Unit of Health Sciences and Technology, University of Oulu, Finland

## Abstract

Chronic back pain (CBP) is a leading cause of work disability worldwide, yet identifying individuals at risk remains difficult due to its multifactorial etiology. This study investigated whether integrating a polygenic risk score (PRS) for CBP with the STarT Back Tool (SBT)—a widely used psychosocial screening instrument—could improve the prediction of work disability, measured as disability leave days over a two-year follow-up. We analysed data from 1,938 participants in the Northern Finland Birth Cohort 1966 with complete genotyping, SBT responses, and registry-linked disability records. A zero-inflated negative binomial regression model was applied to account for the highly skewed distribution of work disability days. Results showed that both SBT and CBP genetic risk independently predicted the cumulative number of disability leave days. While SBT was also associated with the likelihood of having no disability leave, CBP genetic risk was not, suggesting that polygenic risk contributes specifically to the burden of disability among affected individuals. Participants in the highest CBP genetic risk quartile experienced significantly more work disability days, supporting a dose-response relationship. The two tools captured complementary domains: SBT reflected modifiable biopsychosocial risks, while the PRS represented fixed genetic liability. This distinction supports the value of integrating a CBP PRS into existing screening frameworks, particularly for early CBP management.

## INTRODUCTION

Chronic low back pain (CBP) remains a substantial clinical and socioeconomic challenge, ranking as the leading cause of disability worldwide(Ferreira et al., 2023). Musculoskeletal disorders, among which CBP belongs, are the main reason for early exclusion from the labour market due to permanent disability(Finnish Centre for Pensions, 2024). Beyond structural factors like spine pathologies and degenerative conditions, other aspects such as biomechanical stresses, psychological states, and genetic predisposition further complicate CBP(Freidin et al., 2019; Tegeder & Lötsch, 2009). Psychological factors, including stress and anxiety, amplify pain perception and contribute significantly to pain chronicity(Pincus et al., 2002). Depression, metabolic conditions, such as high body mass index (BMI) and diabetes, and smoking are among other important risk factors for CBP(Nieminen et al., 2021). These and other biopsychosocial variables have been integrated into validated models that reliably estimate the risk and progression of chronic pain(Tanguay-Sabourin et al., 2023).

A key challenge in managing low back pain lies in understanding the transition from acute to chronic stages. Although CBP is traditionally defined by its persistence beyond three months, this transition reflects more than just temporal duration — it involves a complex interplay of ongoing nociceptive input, central nervous system plasticity, psychological factors, and individual biological vulnerabilities, including genetic, endocrine, and immune system influences(Zimney et al., 2023). Notably, the heritability of CBP-related disability has been estimated at approximately 40%(Battié et al., 2007). Genome-wide association studies (GWAS) have been instrumental in elucidating the genetic architecture of complex diseases like CBP by identifying genetic variants associated with disease susceptibility. These studies suggest that genetic factors involved in CBP may influence not only pain perception but also broader domains such as psychiatric, sociodemographic and anthropometric traits, including depression, educational attainment and BMI(Freidin et al., 2019; Stanaway et al., 2025).

In primary care settings, the STarT Back Tool (SBT) is widely utilized to stratify acute low back pain patients according to their risk of chronicity(Hill et al., 2008). SBT integrates physical symptoms (pain intensity, functional impairment) and psychosocial factors (anxiety, depression, fear-avoidance beliefs) to categorize patients into low-, medium-, and high-risk groups. This risk stratification aims to guide treatment intensity—from general advice for low-risk patients to psychologically informed physiotherapy for high-risk patients — to prevent the development of CBP(Hill et al., 2008). Despite its international use, evidence of its predictive power is mixed(Karran et al., 2017; Lheureux & Berquin, 2019) and the need to improve the tool(Croft et al., 2024), especially for work-related outcomes, has been identified(Unsgaard-Tøndel et al., 2021).

Polygenic risk scores (PRS), developed from GWAS, aggregate genetic risk variants to estimate individual disease risk. PRS are used for patient stratification, to enhance diagnostic accuracy and to predict outcomes from interventions or assist in subsequent monitoring(Zheutlin & Ross, 2018). For example, in cardiovascular medicine, a recent work showed that integrating ancestry-specific PRS into existing clinical risk models significantly improved prediction of coronary artery disease, particularly in individuals at borderline and intermediate clinical risk — precisely where decision-making is often most uncertain(Ratman et al., 2025).

The primary objective of this population-based cohort study was to investigate whether combining a CBP polygenic risk score with SBT outcomes could improve risk stratification for work disability, specifically measured as disability leave days over a two-year follow-up period using national register data. We hypothesized that integrating genetic risk information would provide complementary insights to the physical and psychosocial dimensions currently assessed in clinical evaluation.

## METHODS

### Study sample

The study sample was drawn from the Northern Finland Birth Cohort 1966 (NFBC1966)(Nordström et al., 2022; University of Oulu, 2024), which consists of individuals born in two provinces of Finland, Oulu and Lapland, whose mothers had expected delivery dates in 1966. The cohort initially included 12,068 pregnant women, resulting in 12,231 children, covering 96.3% of all births in the region that year. Since their mothers’ first antenatal clinic visit, repeated data collections have tracked cohort members longitudinally.

During the 46–year follow-up phase from 2012 to 2014, when the participants were 45–47 years old, those still alive and with known contact details were invited to participate in a health survey, either electronically or by post, and attend a clinical examination. A total of 7,148 participants responded to the questionnaire; of those, 3403 reported having LBP within the past 12 months and responded to the SBT questionnaire. Data from the questionnaire and clinical assessments were then linked to register-based records on sick leave and disability pensions for those who provided written consent.

A total of 5404 participants from NFBC1966 were genotyped using Illumina Human CNV370-Duo DNA bead chip as previously described elsewhere(Sabatti et al., 2009) and imputed using IMPUT2 and 1000 Genomes phase 3 reference panel. Overall, 2112 had both genotype data and responded to the SBT questionnaire. From those, 1938 participants were successfully linked to sick leave and disability pension register data and were not on a disability pension at baseline. Each participant was individually tracked from the registers over a two-year follow-up period starting the day after completing the baseline questionnaire. If this was not known (a participant had not reported it), the clinical examination date was used.

The study followed the principles of the Declaration of Helsinki. The participants took part on a voluntary basis and signed informed consent forms. The Northern Ostrobothnia Hospital District Ethical Committee has approved the study, 94/2011 (12.12.2011).

### Work disability days

The assessment of disability-related days absent from work involved the aggregation of all sickness absence days and disability pension days into a single ‘work disability days’ variable, as used in a prior NFBC1966 study by Varanka-Ruuska et al., 2020(Varanka-Ruuska et al., 2020). Data for this variable were extracted from two national Finnish registers, the Finnish Centre for Pensions(Finnish Centre for Pensions, 2024) (FCP) and the Social Insurance Institution of Finland (SII, Kela register)(The Social Insurance Institution of Finland, n.d.).

To accurately capture the data, it was noted that in Finland, sick leave days are registered into the FCP and Kela registers only after varying deductible times. For the Kela register, only sick leave periods exceeding 10 days (or four days for entrepreneurs) are recorded, while for the FCP register, which primarily includes data on sick leave due to accidents, the first four days are covered by employees and are not recorded. Both full- and part-time sick leave days were considered to be included in the sick leave days outcome. Participants were considered to have a disability pension day if they were granted either a fixed-term or permanent disability pension, regardless of whether it was full-time or part-time. In most cases, to be able to get disability pension benefits in Finland, a person must have been on a sick leave for one year before the eligibility for disability pension is evaluated.

### Polygenic Risk Score (PRS)

Tsepilov et al. (2020) recently developed the PRS specifically for CBP using GWAS data from the UK Biobank cohort(Tsepilov et al., 2023). Briefly, the PRS was derived from a CBP GWAS that used genotype data imputed via IMPUT2 software and the 1000 Genomes phase 3 reference panel from the UK Biobank cohort. Consistent with prior methods, PRS values were standardized using a z-score transformation to facilitate comparison across participants. While the CBP PRS demonstrated moderate predictive ability (AUC = 0.56; 95% CI: 0.56–0.57), its validation across independent cohorts reinforces its potential for clinical application.

### STarT Back Tool

Participants completed a validated Finnish version of the SBT (Piironen et al., 2016). The original version is provided in methods S1 and the translated version is provided in methods S2 (see supplementary materials). SBT scores were calculated by summing positive responses to the nine questionnaire items, resulting in total scores ranging from 0 to 9, with higher scores indicating increased risk of chronic back pain. The nine questions evaluate leg pain referral, comorbid pain in the neck or shoulder, disability in daily activities, pain bothersomeness, and five psychosocial domains: fear, anxiety, catastrophizing, depression, and low confidence in activity.

### Covariates

Sex was recorded at birth, thus dichotomized as female and male. BMI was calculated using weight and height measured at clinical examination. Participants were grouped as ‘non-smokers’, ‘former smokers’, or ‘current smokers’ based on how they had responded to the questions: “Have you ever been a regular smoker?” and “Do you smoke now?” Educational attainment was classified into two levels based on total years of schooling completed by age 46: 12 years or less, including no formal education beyond secondary education; and 13 or more years, representing post-secondary or higher education. Occupational status was obtained from questionnaire and classifies into unemployed, employed (working full-time or part-time, self-employed or entrepreneur, employed/educated by labour market support), and other status such students, on parental/sabbatical leave, homemakers, or other activities not listed before.

### Association analysis

Initial exploratory associations among variables were assessed using Spearman’s correlation. Given the skewed distribution and high frequency of zero counts in work disability days, we first conducted univariate analyses using negative binomial regression to evaluate association with SBT and PRS. To address potential biases from excess zero counts, we subsequently applied a zero-inflated negative binomial regression model. This enabled separate modelling of (a) factors associated with the occurrence (logistic component) and (b) factors predicting the total day number (negative binomial component) of work disability.

Exponentiated coefficients from the negative binomial regression are presented as rate ratios (RRs). The outcome modelled, total work disability days, is technically a duration measure rather than a discrete event count, the negative binomial framework models it as count data for statistical purposes. Using the term ‘incidence rate ratio’ (IRR) would be misleading in this context, as our outcome does not reflect new event occurrences over person-time. Therefore, we refer to the exponentiated estimates as ‘rate ratios’ (RRs) to more accurately describe the multiplicative change in expected work disability days associated with a one-unit change in each predictor variable. For the logistic component of the zero-inflated model, results are presented as odds ratios (ORs), representing the odds of having no disability leave days (i.e., belonging to the structural zero group) associated with each predictor. An odds ratio greater than 1 indicates higher odds of no disability leave, while an odds ratio less than 1 indicates lower odds. Confidence intervals (95% CIs) for both RRs and ORs were calculated using the standard normal approximation, based on ±1.96 times the standard error of the coefficient estimates.

Zero-inflated regression analyses were performed using the zeroinfl function from the pscl package in R, specifying negative binomial distribution for the count portion. Covariates included in all multivariate regression analyses were sex, BMI, smoking status (never, former, or current smoker), education level (up to secondary education or post-secondary education), and occupational status (employed, unemployed, or other). Smoking status, occupational status, sex and educational level were considered categorical variables; the remaining variables including work disability days, SBT, PRS and BMI were treated as continuous variables.

To explore possible non-linear effects, participants were categorized into PRS quartiles, allowing investigation of disability leave days across different levels of genetic risk. All analyses were performed using R statistical software. Statistical significance was defined at the conventional threshold of p < 0.05.

To visualise the effect of SBT and PRS risk groups on the total number of work disability days, the expected work disability days were calculated at the different values of those variables, keeping all other confounders constant and using the negative binomial estimates of the zero-inflated regression. In particular, expected work disability days at different SBT and PRS risk groups were calculated for an employed non-smoker male with the sample mean BMI and no higher education. Predicted values were obtained by setting the covariates to these reference levels and varying SBT and PRS categories accordingly, using the fitted model to compute marginal means.

## RESULTS

### Study sample and main data

Among the 1,938 NFBC1966 subjects with available SBT, PRS, and leave days data, 1,713 had complete information on sex, BMI, smoking status, education level, and occupational status. The descriptors are summarised in Table 1.

**Table 1.** Table presenting the main descriptors of the sample characteristics including main confounder variables.

| Category | Statistics |
| --- | --- |
| **Sample (complete data)** | 1,713 subjects, 46 years old, 56.3% female |
| **Work disability days (all subjects)** | 0–730 days; 75.6% had no disability leave days |
| **Work disability days (with disability)** | median 67.95 days (IQR 61.50) |
| **SBT score** | 0–9 range; mean $1.6 \pm 1.6$ |
| **Chronic back pain PRS** | -3.32 to 3.47 (z-score normalized) |
| **BMI (mean ± SD)** | $26.8 \pm 4.8$ |
| **Smoking status** | 47% never, 30% former, 23% current smokers |
| **Education (post-secondary)** | 25% |
| **Employment** | 91% employed, 4% unemployed, 5% other status |

Mean age was approximately 46 years at the time of sampling. The sample was 56.3% female, with a mean BMI of 26.8 kg/m^2^ (SD = 4.8). Of the sample having complete data, 47% had never smoked and 23% were current smokers. Seventy-five percent of the sample had up to secondary education, and only 4% of the participants were unemployed at the time of the questionnaire. Work disability ranged from 0 to 730 days, with 75.6% presenting no work disability accumulated over two-year period. Subjects with work disability had median of 67.95 days and an inter-quartile range of 61.50 days. SBT questionnaire responses were summed to yield a score ranging from 0 to 9, with a mean of 1.6 (SD = 1.6). The CBP PRS was normally distributed and normalized using a z-score, ranging from -3.32 to 3.47. Distributions of disability leave days, SBT summed scores, and PRS scores are shown in Figure S1 (supplementary materials).

### Univariate analysis

In the univariate analysis, higher SBT scores were correlated with greater work disability days (Spearman’s correlation coefficient ρ = 0.185, *p* = 2.14 × 10^-17^; Figure 1A). In univariate regression, increased SBT scores were associated with a higher total number of work disability days and a lower likelihood of having no disability leave (RR = 1.17, 95% CI: 1.08–1.28; OR = 0.78, 95% CI: 0.73–0.85). In contrast, CBP PRS was not significantly correlated with work disability days (Spearman’s ρ = 0.015, *p* = 0.54; Figure 1B). Regression analysis showed that genetic risk was not associated with the odds of having no work disability (OR = 0.98, 95% CI: 0.87–1.11), although among individuals with disability, higher genetic risk was associated with a greater total number of work disability days (RR = 1.35, 95% CI: 1.15–1.60). Additionally, CBP PRS was weakly but significantly correlated with higher SBT scores (ρ = 0.08, *p* = 0.002; Figure 1C) and with higher SBT-predicted risk group classifications (low, medium, and high risk; ρ = 0.070, *p* = 0.004; Figure 1D).

**Figure 1.**
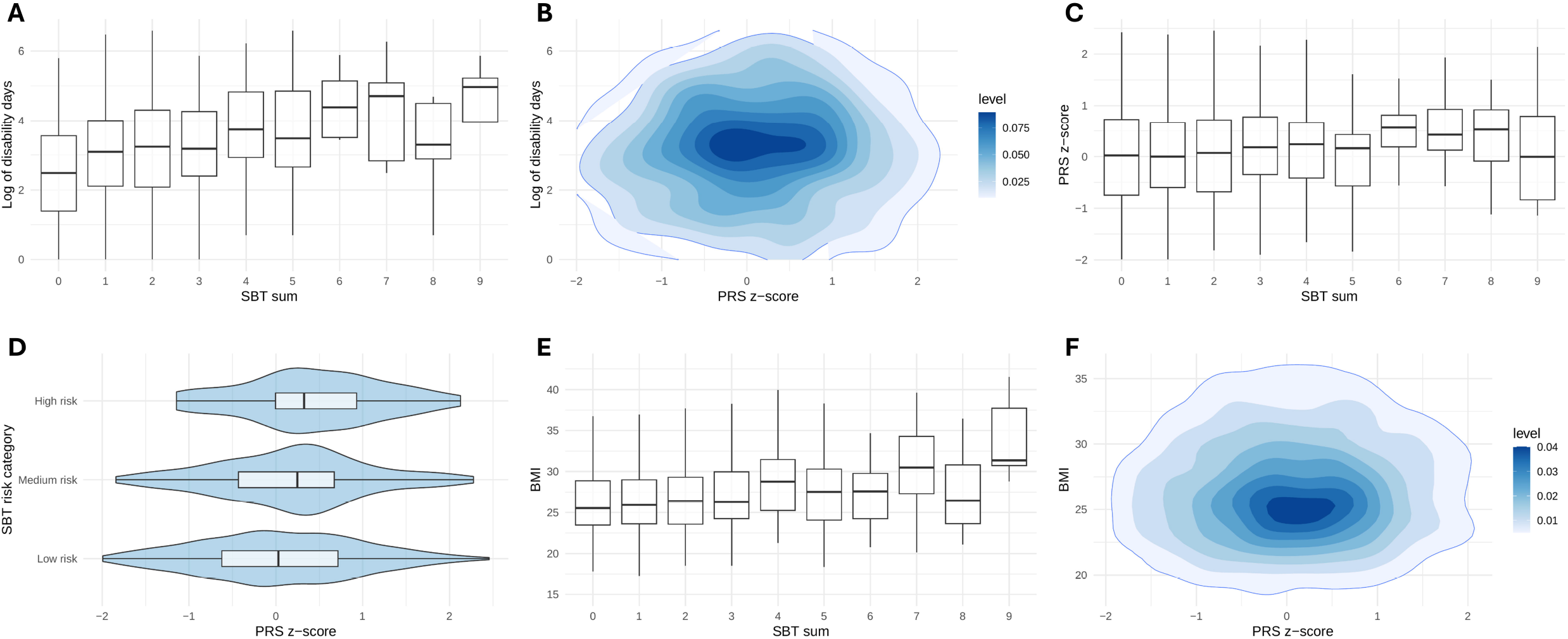
Pairwise correlations among variables. Distributions of disability leave days by SBT scores and PRS (A, B); distribution of PRS by SBT scores and SBT-defined chronicity risk groups (C, D); distribution of BMI by SBT scores and PRS (E, F).

Higher SBT scores were also correlated with increased BMI (ρ = 0.12, *p* = 5.847 × 10^-17^; Figure 1E), while no significant correlation was found between PRS and BMI (ρ = 0.03, *p* = 0.16; Figure 1F). SBT was further correlated with lower education level (ρ = –0.16, *p* < 0.001), unemployed occupational status (ρ = –0.10, *p* < 0.001), female sex (ρ = 0.067, *p* = 0.006), and smoking status (ρ = 0.09, *p* = 1.81 × 10^-4^). In contrast, the PRS was not significantly correlated with sex (ρ = 0.023, *p* = 0.35) or education level (ρ = –0.012, *p* = 0.61), but showed weak correlations with unemployed occupational status (ρ = –0.052, *p* = 0.033) and smoking status (ρ = 0.07, *p* = 0.008).

### Multivariate Analysis: Zero-Inflated Model

In multivariate analysis, SBT was a significant predictor in both components of the model similarly to univariate analysis. Higher SBT scores were associated with a longer cumulative duration of work disability (RR = 1.16, 95% CI: 1.08–1.26) and individuals with higher SBT scores were less likely to present no disability (OR = 0.81, 95% CI: 0.75–0.87). Likewise, higher PRS scores were also associated with an increased total number of work disability days (RR = 1.33, 95% CI: 1.17–1.51) but not with changes in likelihood of no disability. Never smokers and employed individuals were associated with lower number of work disability days, compared to current smokers and unemployed, respectively. Higher BMI was slightly but significantly associated with lower odds of having no work disability (OR = 0.96, 95% CI: 0.94–0.99), but among individuals who did take work disability days, it was also associated with a modest reduction in the total number of work disability days (RR = 0.96, 95% CI: 0.94–0.98). Additionally, males were more likely of no taking work disability than females. The effect estimates and statistical significance of all predictors from the adjusted zero-inflated regression are summarized in Table 2.

**Table 2.** Statistics from the zero-inflated regression of work disability days using SBT, PRS as predictors including adjustments for sex, smoking status, employment status and BMI. BMI: body mass index; PRS: polygenic risk score, SBT: STarT Back Tool; RR: rate ratio; OR: odds ratio; CI: confidence intervals.

**Negative binomial regression predicting total work disability days (count distribution)**
|  | RR | 95% CI |  | p-value |
| --- | --- | --- | --- | --- |
| **SBT score** | 1.16 | 1.08 | 1.26 | 0.000 |
| **PRS** | 1.33 | 1.17 | 1.51 | 0.000 |
| **Sex (male)** | 0.80 | 0.60 | 1.06 | 0.124 |
| **Ex-smoker (vs current smoker)** | 1.14 | 0.80 | 1.64 | 0.458 |
| **Never smoked (vs current smoker)** | 0.61 | 0.43 | 0.85 | 0.004 |
| **Post-secondary education** | 1.34 | 0.95 | 1.89 | 0.092 |
| **Employed (vs unemployed)** | 0.32 | 0.16 | 0.65 | 0.002 |
| **Other employed status (vs unemployed)** | 0.82 | 0.45 | 1.49 | 0.521 |
| **BMI** | 0.96 | 0.94 | 0.99 | 0.016 |
| **Dispersion parameter estimate ($\theta$)** | 0.49 | 0.40 | 0.60 | 0.000 |

**Logistic regression predicting probability of no disability days (structural zeros)**
|  | OR | 95% CI |  | p-value |
| --- | --- | --- | --- | --- |
| **SBT score** | 0.81 | 0.75 | 0.87 | 0.000 |
| **PRS** | 1.04 | 0.92 | 1.17 | 0.544 |
| **Sex (male)** | 1.40 | 1.10 | 1.79 | 0.007 |
| **Ex-smoker (vs current smoker)** | 1.03 | 0.74 | 1.42 | 0.840 |
| **Never smoked (vs current smoker)** | 1.28 | 0.95 | 1.74 | 0.109 |
| **Post-secondary education** | 1.29 | 0.96 | 1.73 | 0.086 |
| **Employed (vs unemployed)** | 0.71 | 0.38 | 1.33 | 0.282 |
| **Other employed status (vs unemployed)** | 1.23 | 0.53 | 2.88 | 0.540 |
| **BMI** | 0.96 | 0.94 | 0.98 | 0.001 |

To investigate whether the PRS exhibits non-linear behaviour, the PRS was categorized into four risk groups based on quantiles within the sample. Subsequently, these four genetic risk groups for back pain were utilized in place of the continuous PRS (multivariate adjusted model). Analysis of the total number of work disability days revealed statistically significant differences between the low-risk and high-risk groups (RR = 1.86, 95% CI: 1.25–2.76), with the effect on work disability days increasing in conjunction with genetic risk. No relevant changes were observed in the SBT association (RR = 1.16, 95% CI: 1.07–1.27, OR = 0.81, 95% CI: 0.75–0.87, see Table S1 in supplementary materials for the statistics of all variables). The relationship between expected mean work disability days and the four genetic risk groups, along with the SBT scores, are illustrated in Figure 2. The expected work disability days nominally increased from low-to high-genetic-risk CBP.

**Figure 2.**
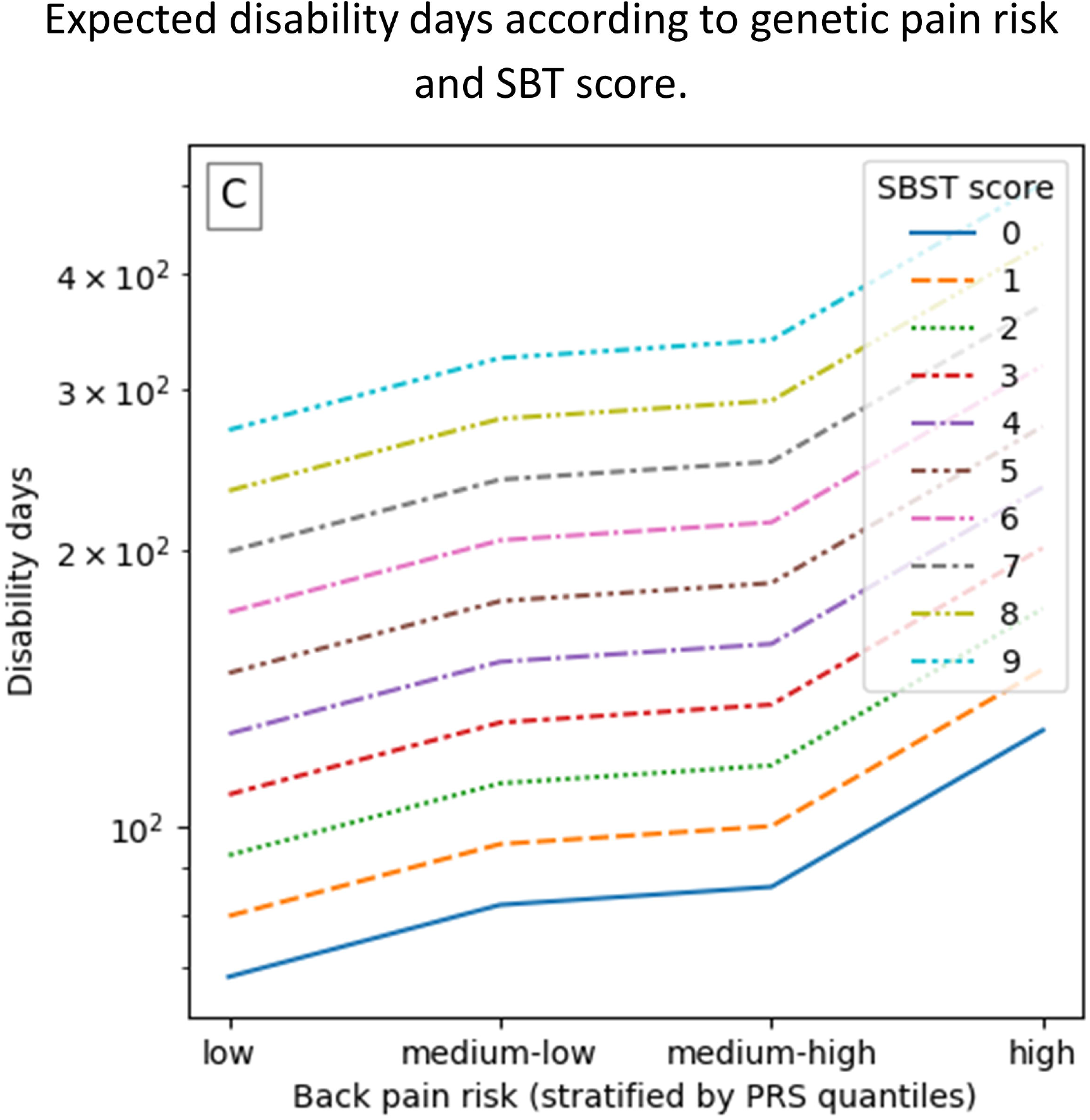
Predicted work disability days based on regression estimates. Expected disability days (log scale, y-axis) across genetic risk groups (x-axis, stratified by PRS quantiles), with lines representing SBST scores from 0 to 9.

## DISCUSSION

CBP remains a significant clinical and socioeconomic burden, underscoring the need for accurate early risk stratification tools to guide management and reduce disability. This study demonstrates that integrating a PRS for CBP with the established SBT improves the characterization of work disability outcomes, specifically measured as the number of work disability days over a two-year period.

The PRS was a significant predictor of work disability total days in both univariate and multivariate zero-inflated models (RR = 1.35, 95% CI: 1.15–1.60; RR = 1.33, 95% CI: 1.17–1.51, respectively), despite showing no significant overall association with the occurrence of work disability. A clear dose-response relationship was observed, with individuals in the highest polygenic risk group experiencing significantly greater numbers of work disability days. In contrast, the SBT score was overall significantly correlated with work disability days (Spearman’s coefficient = 0.185, *p* = 2.14 × 10^-17^), and consistently associated with disability outcomes in both univariate and multivariate analyses. Higher SBT scores were robustly linked to higher number of work disability days (RR = 1.17, 95% CI: 1.08–1.28; RR = 1.16, 95% CI: 1.08–1.26, respectively) and to lower odds of having no work disability days (OR = 0.78, 95% CI: 0.73–0.85; OR = 0.81, 95% CI: 0.75–0.87, respectively). These findings highlight the potential clinical utility of incorporating genetic risk information alongside established screening tools to enhance early risk stratification and guide targeted interventions for chronic back pain.

Our use of zero-inflated negative binomial regression allowed us to disentangle predictors of the likelihood of experiencing any work disability (logistic component) from those affecting its total cumulative duration (count component). Notably, genetic risk was only associated with the severity (i.e., number of work disability days) of disability, not its onset. This distinction suggests that genetic predisposition may play a larger role in determining the chronicity of back pain rather than the initial occurrence of disability. Conversely, SBT captures both functional and psychosocial aspects that influence both risk and persistence of disability. Together, these patterns support the view that PRS and SBT tap into distinct but complementary domains of vulnerability such as biological versus biopsychosocial respectively.

This interpretation is further reinforced by the finding that SBT scores were significantly correlated with BMI—a well-established metabolic and physical risk factor for disability— whereas PRS showed no such relationship. More broadly, SBT demonstrated consistent correlations with several modifiable sociodemographic and behavioral factors, including education, occupational status, and smoking, as well as non-modifiable traits like sex. In contrast, PRS was largely uncorrelated with these variables, showing only weak associations with occupational status and smoking. Nevertheless, the correlation of back pain genetic risk with smoking is not unexpected, as genetic correlation between back pain and smoking has already been reported(Freidin et al., 2019). Altogether, these differences highlight how the two measures complement each other: SBT reflects mainly changeable, real-world factors influencing disability —and probably pain—, while PRS captures mostly fixed genetic risk.

Stratifying individuals by PRS quartiles further supported its predictive value: those in the highest genetic risk group experienced significantly more work disability days, consistent with a dose-response relationship. While PRS had no impact on the likelihood of avoiding disability, it effectively predicted the burden among those who were affected — a pattern consistent with SBT’s role in early care settings, where symptoms have already emerged. Importantly, these findings highlight the potential for PRS to enrich existing screening approaches at population-level. Adding genetic data to clinical tools like SBT could help identify people at risk of prolonged work disability. Naturally, further studies in patient samples are needed but this integration may have a potential to support more personalized treatment planning, including earlier referrals for intensive interventions among those genetically predisposed to worse outcomes.

In addition to clinical tools and genetic predisposition, our study identified modifiable predictors associated with work disability outcomes. Never smoking and being employed were linked to a lower number of work disability days, emphasizing the protective effects of healthier lifestyle behaviours. Although higher BMI was associated with an increased likelihood of experiencing work disability, it was also linked to a modest reduction in the total duration of the work disability among those affected. This nuanced finding suggests that while BMI may influence the risk of disability occurrence, it has a more complex or less straightforward relationship with the severity of work disability once it occurs. One speculative explanation is that occupational health practices in Finland, such as the implementation of alternative duty work policies, may be more frequently applied to individuals with obesity, given that obesity is a well-recognized and visible risk factor for reduced work ability. These policies allow employees to perform modified duties instead of taking sick leave, which may contribute to a shorter duration of work disability in this group. Additionally, males showed a higher probability of not registering disability leave compared to females. These findings highlight the important role of modifiable social and behavioural factors in shaping disability outcomes and suggest potential targets for intervention alongside clinical and genetic risk stratification.

The strengths of our study include the use of objectively recorded disability data from national registries, a large and representative population-based cohort, and the application of advanced modelling techniques that account for the zero-inflated nature of work disability days data. To our knowledge, this is the first study to empirically test and demonstrate the value of integrating a validated PRS for back pain with the SBT tool, offering early evidence that such integration may meaningfully improve risk stratification in primary care.

Nonetheless, certain limitations must be acknowledged. First, the outcome measure — total work disability days — did not distinguish between causes of disability, meaning that not all disability days may have been directly attributable to back pain. Future studies incorporating diagnostic codes (e.g., ICD-10 or ICD-11) could provide more specific back pain-related estimates. However, because CBP is a major contributor to work disability, the use of a more specific outcome would likely enhance association power and further strengthen the findings reported here. Second, the observational nature of the study limits causal inference. Although we adjusted for major confounders, the possibility of residual confounding or unmeasured variables cannot be excluded. Finally, although the SBT captures biopsychosocial factors, it does not explicitly include occupational or work-related determinants, which may also influence disability outcomes(Unsgaard-Tøndel et al., 2021).

In conclusion, this study provides novel evidence that integrating genetic risk information with clinical screening improves the ability to identify individuals at higher risk of disability due to CBP. While future studies are needed to refine prediction models and validate these findings across broader populations and outcome measures, this work lays an important foundation for the use of genetic risk stratification for back pain in primary care. Incorporating PRS into early back pain assessments may support more targeted management strategies, ultimately reducing the personal and societal burden of CBP.

## ACKNOWLEDGEMENTS

We wish to thank all cohort members, researchers and NFBC project center personnel who participated in the NFBC data collections. This project was supported by the Marie Skłodowska Curie International Training Network (ITN) “disc4all”(https://disc4all.upf.edu, accessed on 1^st^, April 2025) grant agreement #955735. NFBC1966 46y follow-up study received financial support from University of Oulu Grant no. 24000692, Oulu University Hospital Grant no. 24301140, ERDF European Regional Development Fund Grant no. 539/2010 A31592. Generative AI (ChatGPT, OpenAI) was used solely to improve the clarity and phrasing of the manuscript. The authors declare no conflicts of interest relevant to the content of this manuscript.

## DATA AVAILABILITY STATEMENT

NFBC data are available from the University of Oulu, Infrastructure for Population Studies. Permission to use the data can be applied for research purposes via an electronic material request portal. In the use of data, we follow the EU general data protection regulation (679/2016) and the Finnish Data Protection Act. The use of personal data is based on a cohort participant’s written informed consent in their latest follow-up study, which may cause limitations to its use. Please, contact the NFBC project center and visit the cohort website (www.oulu.fi/nfbc) for more information.

## AUTHOR CONTRIBUTIONS

This study was designed by R.C., M.K.N. and F.M.K.W. The data were analyzed by R.C., M.K.N., E.H., and T.M. The results were critically examined by all authors. R.C. had a primary role in preparing the manuscript, which was reviewed and edited by J.K. and F.M.K.W. All authors have approved the final version of the manuscript and agree to be accountable for all aspects of the work.

## REFERENCES

Battié, M. C., Videman, T., Levalahti, E., Gill, K., & Kaprio, J. (2007). Heritability of low back pain and the role of disc degeneration. Pain, 131(3), 272–280. 10.1016/j.pain.2007.01.010

Croft, P., Hill, J. C., Foster, N. E., Dunn, K. M., & van der Windt, D. A. (2024). Stratified health care for low back pain using the STarT Back approach: Holy Grail or doomed to fail? Pain. 10.1097/J.PAIN.0000000000003319

Ferreira, M. L., De Luca, K., Haile, L. M., Steinmetz, J. D., Culbreth, G. T., Cross, M., Kopec, J. A., Ferreira, P. H., Blyth, F. M., Buchbinder, R., Hartvigsen, J., Wu, A. M., Safiri, S., Woolf, A. D., Collins, G. S., Ong, K. L., Vollset, S. E., Smith, A. E., Cruz, J. A., … March, L. M. (2023). Global, regional, and national burden of low back pain, 1990–2020, its attributable risk factors, and projections to 2050: a systematic analysis of the Global Burden of Disease Study 2021. The Lancet Rheumatology, 5(6), e316–e329. 10.1016/S2665-9913(23)00098-X

Finnish Centre for Pensions. (2024). Statistics. https://www.etk.fi/en/research-statistics-and-projections/statistics/

Freidin, M. B., Tsepilov, Y. A., Palmer, M., Karssen, L. C., Suri, P., Aulchenko, Y. S., & Williams, F. M. K. (2019). Insight into the genetic architecture of back pain and its risk factors from a study of 509,000 individuals. Pain, 160(6), 1361–1373. 10.1097/j.pain.0000000000001514

Hill, J. C., Dunn, K. M., Lewis, M., Mullis, R., Main, C. J., Foster, N. E., & Hay, E. M. (2008). A primary care back pain screening tool: Identifying patient subgroups for initial treatment. Arthritis Care and Research, 59(5), 632–641. 10.1002/art.23563

Karran, E. L., McAuley, J. H., Traeger, A. C., Hillier, S. L., Grabherr, L., Russek, L. N., & Moseley, G. L. (2017). Can screening instruments accurately determine poor outcome risk in adults with recent onset low back pain? A systematic review and meta-analysis. BMC Medicine, 15(1), 1–15. 10.1186/S12916-016-0774-4/TABLES/8

Lheureux, A., & Berquin, A. (2019). Comparison between the STarT Back Screening Tool and the Örebro Musculoskeletal Pain Screening Questionnaire: Which tool for what purpose? A semi-systematic review. Annals of Physical and Rehabilitation Medicine, 62(3), 178–188. 10.1016/J.REHAB.2018.09.007

Nieminen, L. K., Pyysalo, L. M., & Kankaanpää, M. J. (2021). Prognostic factors for pain chronicity in low back pain: A systematic review. Pain Reports, 6(1). 10.1097/PR9.0000000000000919

Nordström, T., Miettunen, J., Auvinen, J., Ala-Mursula, L., Keinänen-Kiukaanniemi, S., Veijola, J., Järvelin, M. R., Sebert, S., & Männikkö, M. (2022). Cohort Profile: 46 years of follow-up of the Northern Finland Birth Cohort 1966 (NFBC1966). International Journal of Epidemiology, 50(6), 1786–1787J. 10.1093/IJE/DYAB109

Piironen, S., Paananen, M., Haapea, M., Hupli, M., Zitting, P., Ryynänen, K., Takala, E. P., Korniloff, K., Hill, J. C., Häkkinen, A., & Karppinen, J. (2016). Transcultural adaption and psychometric properties of the STarT Back Screening Tool among Finnish low back pain patients. European Spine Journal, 25(1), 287–295. 10.1007/S00586-015-3804-6/TABLES/4

Pincus, T., Kim Burton, A., Vogel, S., & Field, A. P. (2002). A Systematic Review of Psychological Factors as Predictors of Chronicity/Disability in Prospective Cohorts of Low Back Pain. SPINE, 27(5), 109–120.

Ratman, D., Tshiaba, P., Levin, M., Sun, J., Tunstall, T., Maier, R., Shah, P., Rabinowitz, M., Rader, D. J., Kumar, A., & Im, K. (2025). Polygenic risk scores improve CAD risk prediction in individuals at borderline and intermediate clinical risk. Npj Cardiovascular Health 2025 2:1, 2(1), 1–14. 10.1038/s44325-025-00049-7

Sabatti, C., Service, S. K., Hartikainen, A. L., Pouta, A., Ripatti, S., Brodsky, J., Jones, C. G., Zaitlen, N. A., Varilo, T., Kaakinen, M., Sovio, U., Ruokonen, A., Laitinen, J., Jakkula, E., Coin, L., Hoggart, C., Collins, A., Turunen, H., Gabriel, S., … Peltonen, L. (2009). Genome-wide association analysis of metabolic traits in a birth cohort from a founder population. Nature Genetics, 41(1), 35–46. 10.1038/NG.271

Stanaway, I. B., Suri, P., Afari, N., Dochtermann, D., Gerstenberger, A., Pyarajan, S., Rosen, E. J., & Gasperi, M. (2025). Multi-ancestry meta-analysis of genome-wide association studies discovers 67 new loci associated with chronic back pain. Nature Communications 2025 16:1, 16(1), 1–12. 10.1038/s41467-024-55326-3

Tanguay-Sabourin, C., Fillingim, M., Guglietti, G. V., Zare, A., Parisien, M., Norman, J., Sweatman, H., Da-ano, R., Heikkala, E., Breitner, J. C. S., Menes, J., Poirier, J., Tremblay-Mercier, J., Perez, J., Karppinen, J., Villeneuve, S., Thompson, S. J., Martel, M. O., Roy, M., … Vachon-Presseau, E. (2023). A prognostic risk score for development and spread of chronic pain. Nature Medicine, 29(7), 1821–1831. 10.1038/s41591-023-02430-4

Tegeder, I., & Lötsch, J. (2009). Current evidence for a modulation of low back pain by human genetic variants. Journal of Cellular and Molecular Medicine, 13(8b), 1605–1619. 10.1111/J.1582-4934.2009.00703.X

The Social Insurance Institution of Finland. (n.d.). Statistics. 2024. https://www.kela.fi/kelan-tutkimus-ja-tilastot

Tsepilov, Y. A., Elgaeva, E. E., Nostaeva, A. V., Compte, R., Kuznetsov, I. A., Karssen, L. C., Freidin, M. B., Suri, P., Williams, F. M. K., & Aulchenko, Y. S. (2023). Development and Replication of a Genome-Wide Polygenic Risk Score for Chronic Back Pain. Journal of Personalized Medicine, 13(6), 977. 10.3390/jpm13060977

University of Oulu. (2024, April). Northern Finland Birth Cohort 1966. University of Oulu. http://urn.fi/urn:nbn:fi:att:bc1e5408-980e-4a62-b899-43bec3755243

Unsgaard-Tøndel, M., Vasseljen, O., Nilsen, T. I. L., Myhre, G., Robinson, H. S., & Meisingset, I. (2021). Prognostic ability of STarT Back Screening Tool combined with work-related factors in patients with low back pain in primary care: a prospective study. BMJ Open, 11(6), e046446. 10.1136/BMJOPEN-2020-046446

Varanka-Ruuska, T., Tolvanen, M., Vaaramo, E., Keinänen-Kiukaanniemi, S., Sebert, S., Rautio, N., & Ala-Mursula, L. (2020). Glucose metabolism in midlife predicts participation in working life: A Northern Finland Birth Cohort 1966 study. Occupational and Environmental Medicine, 77(5), 324–332. 10.1136/oemed-2019-106170

Zheutlin, A. B., & Ross, D. A. (2018). Polygenic Risk Scores: What Are They Good For? Biological Psychiatry, 83(11), e51–e53. 10.1016/J.BIOPSYCH.2018.04.007

Zimney, K., Van Bogaert, W., & Louw, A. (2023). The Biology of Chronic Pain and Its Implications for Pain Neuroscience Education: State of the Art. Journal of Clinical Medicine 2023, Vol. 12, Page 4199, 12(13), 4199. 10.3390/JCM12134199

