## supplementary materials for "Does combining the STarT Back Tool with a polygenic risk score for chronic low back pain improve prediction of work disability over two years?"

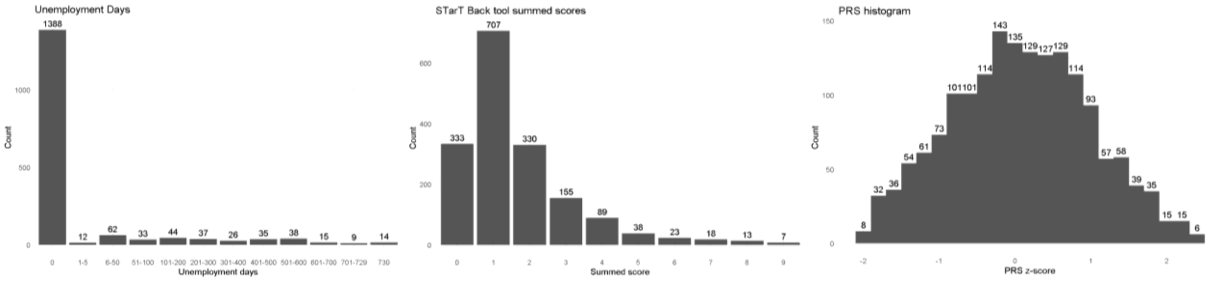


*Figure S1. Distributions of the main variables. Disability leave days (left); STarT Back Tool summed score (SBT, middle); chronic back pain polygenic risk score (PRS, right).*

*Table S1. Statistic of the regression of disability days using stratified PRS by quantiles in four risk groups. BMI: body mass index; PRS: polygenic risk score, SBT: STarT Back Tool; RR: rate ratio; OR: odds ratio; CI: confidence intervals.*

**Negative binomial regression predicting total disability leave days (count distribution)**

| | RR | 95% CI | | p-value |
|---|---|---|---|---|
| **SBT score** | 1.16 | 1.07 | 1.27 | 0.001 |
| **PRS risk medium-low (vs low)** | 1.20 | 0.80 | 1.80 | 0.384 |
| **PRS risk medium-high (vs low)** | 1.25 | 0.84 | 1.86 | 0.301 |
| **PRS risk high (vs low)** | 1.86 | 1.25 | 2.76 | 0.002 |
| **Sex (male)** | 0.83 | 0.62 | 1.11 | 0.200 |
| **Ex-smoker (vs current smoker)** | 1.16 | 0.80 | 1.67 | 0.429 |
| **Never smoked (vs current smoker)** | 1.23 | 0.87 | 1.73 | 0.234 |
| **Post-secondary education** | 0.73 | 0.51 | 1.03 | 0.075 |
| **Employed (vs unemployed)** | 1.41 | 0.50 | 3.94 | 0.464 |
| **Other employed status (vs unemployed)** | 0.97 | 0.94 | 1.00 | 0.513 |
| **BMI** | 0.47 | 0.38 | 0.58 | 0.028 |
| **Dispersion parameter estimate (θ)** | 1.16 | 1.07 | 1.27 | 0.000 |

**Logistic regression predicting probability of no disability days (structural zeros)**

| | OR | 95% CI | | p-value |
|---|---|---|---|---|
| **SBT score** | 0.81 | 0.75 | 0.87 | 0.000 |
| **PRS risk medium-low (vs low)** | 0.98 | 0.69 | 1.39 | 0.921 |
| **PRS risk medium-high (vs low)** | 1.14 | 0.80 | 1.61 | 0.471 |
| **PRS risk high (vs low)** | 1.06 | 0.77 | 1.46 | 0.711 |
| **Sex (male)** | 1.40 | 1.10 | 1.79 | 0.007 |
| **Ex-smoker (vs current smoker)** | 1.03 | 0.75 | 1.43 | 0.835 |
| **Never smoked (vs current smoker)** | 1.28 | 0.95 | 1.74 | 0.109 |
| **Post-secondary education** | 0.78 | 0.55 | 1.11 | 0.165 |
| **Employed (vs unemployed)** | 0.71 | 0.38 | 1.32 | 0.278 |
| **Other employed status (vs unemployed)** | 1.29 | 0.55 | 3.02 | 0.563 |
| **BMI** | 0.96 | 0.93 | 0.98 | 0.001 |

*Methods S1. The original English version of the STarT Back Tool (SBT) questionnaire.*

### The Keele STarT Back Screening Tool

Patient name: ________________________________ Date: _____________

Thinking about the **last 2 weeks** tick your response to the following questions:

| | | Disagree 0 | Agree 1 |
|---|---|---|---|
| 1 | My back pain has **spread down my leg(s)** at some time in the last 2 weeks | ☐ | ☐ |
| 2 | I have had pain in the **shoulder** or **neck** at some time in the last 2 weeks | ☐ | ☐ |
| 3 | I have only **walked short distances** because of my back pain | ☐ | ☐ |
| 4 | In the last 2 weeks, I have **dressed more slowly** than usual because of back pain | ☐ | ☐ |
| 5 | It's not really safe for a person with a condition like mine to be physically active | ☐ | ☐ |
| 6 | **Worrying thoughts** have been going through my mind a lot of the time | ☐ | ☐ |
| 7 | I feel that **my back pain is terrible** and **it's never going to get any better** | ☐ | ☐ |
| 8 | In general I have **not enjoyed** all the things I used to enjoy | ☐ | ☐ |

9. Overall, how **bothersome** has your back pain been in the **last 2 weeks**?

| Not at all | Slightly | Moderately | Very much | Extremely |
|---|---|---|---|---|
| ☐ | ☐ | ☐ | ☐ | ☐ |
| 0 | 0 | 0 | 1 | 1 |

**Total score (all 9):** __________________ **Sub Score (Q5-9):**_____________

© Keele University 01/08/07
Funded by Arthritis Research UK

*Methods S2. Finnish version of SBT.*

### Keele STarT selkäkysely

Tutkittavan nimi: ______________________________ Päiväys: _____________

Ajattele **viimeksi kulunutta 2 viikkoa** vastatessasi seuraaviin kysymyksiin:

| | | **Eri mieltä** 0 | **Samaa mieltä** 1 |
|---|---|---|---|
| 1 | Selkäkipuni on **säteillyt alaraajaani (-raajoihini)** jossakin vaiheessa viimeksi kuluneen 2 viikon aikana | ☐ | ☐ |
| 2 | Minulla on ollut **niska-** tai **hartiakipua** jossakin vaiheessa viimeksi kuluneen 2 viikon aikana | ☐ | ☐ |
| 3 | Olen **kävellyt ainoastaan lyhyitä matkoja** selkäkipuni vuoksi | ☐ | ☐ |
| 4 | Viimeksi kuluneen 2 viikon aikana **pukeutumiseni on ollut tavallista hitaampaa** selkäkivun vuoksi | ☐ | ☐ |
| 5 | Tällaisessa kunnossa olevan henkilön ei ole oikeastaan turvallista olla fyysisesti aktiivinen (harrastaa liikuntaa, työskennellä) | ☐ | ☐ |
| 6 | Olen ollut usein **huolestunut tilanteestani** | ☐ | ☐ |
| 7 | Minusta tuntuu, että **selkäkipuni on erittäin vaikea** eikä **se tule koskaan paremmaksi** | ☐ | ☐ |
| 8 | Yleisesti ottaen **en ole nauttinut** kaikista niistä asioista, joista ennen nautin | ☐ | ☐ |

9. Kuinka **haittaavaa** selkäkipusi on ollut **viimeksi kuluneen 2 viikon** aikana?

| Ei lainkaan | Hieman | Kohtalaisesti | Paljon | Erittäin paljon |
|---|---|---|---|---|
| ☐ 0 | ☐ 0 | ☐ 0 | ☐ 1 | ☐ 1 |

**Kokonaispisteet (kaikki 9):** ___________ **Osapisteet (kysymykset 5-9):**___________

© Keele University 01/08/07
Funded by Arthritis Research UK
